# Complex associations of genetic/epigenetic variations of CGG repeats with patient phenotypes in oculopharyngodistal myopathy

**DOI:** 10.1101/2025.05.13.25327490

**Authors:** Nobuyuki Eura, Satoru Noguchi, Megumu Ogawa, Kyuto Sonehara, Ai Yamanaka, Shinichiro Hayashi, Yukinori Okada, Kazuma Sugie, Ichizo Nishino

## Abstract

Oculopharyngodistal myopathy (OPDM) is caused by CGG triplet repeat expansions in six genes. To explore the genetics and epigenetics of OPDM, we conducted CRISPR/Cas9-targeted resequencing of repeat regions in 89 patients. Repeat regions essentially comprised pure CGG expansions, but exhibited size variability, even within patients. Expanded *LRP12* and *GIPC1* alleles showed distinct single nucleotide variant patterns, suggesting founder haplotypes. *LRP12*-expanded reads lacked flanking sequences present in non-expanded reads, whereas *GIPC1* expanded repeats contained specific nucleotide patterns in their 5’-regions. Structural variations were identified in some patients. A significant inverse correlation was observed between repeat length and age at onset in patients with *GIPC1* or *NOTCH2NLC* expansions, while this was disturbed by higher methylation of expanded regions in patients with *LRP12* expansions, leading to delayed onset. These findings reveal a complex interplay among repeat size, sequence context, and epigenetic state in OPDM pathogenesis, advancing knowledge and providing opportunities for therapeutic intervention.

## Introduction

Oculopharyngodistal myopathy (OPDM) is an autosomal dominant hereditary muscle disease characterized by slowly progressive ptosis, ophthalmoplegia, bulbar weakness, and muscle weakness predominantly of the distal limbs, with pathological features including small angular fibers and rimmed vacuoles. OPDM etiology has been linked to CGG repeat expansions in the 5’-untranslated regions of several genes, including: *LRP12*, *GIPC1*, *NOTCH2NLC*, *RILPL1*, *LOC642361/NUTM2B-AS1*, and *ABCD3*^1–7^. Despite their distinct genetic origins, patients with different OPDM subtypes exhibit similar clinicopathological features, implying shared underlying pathogenic features; however, the precise disease mechanism remains unclear, with two major hypotheses proposed: (1) protein toxicity caused by repeat-associated non-AUG (RAN) translation, and (2) RNA toxicity due to RNA foci formation disrupting RNA metabolism^8^. Additionally, some asymptomatic carriers exhibit extensive repeat expansions, with hypermethylation of the expanded regions, suggesting that epigenetic modifications, such as promoter hypermethylation, may prevent disease development ^2, 4, 9,10^. Thus, elucidating the sequence composition and epigenetic landscape of expanded repeats is crucial for understanding OPDM pathogenesis.

Genetic diagnosis of OPDM traditionally relies on repeat-primed PCR (RP-PCR), amplicon-length analysis PCR (AL-PCR), and Southern blotting^1–3, 10^; these methods detect repeat expansions, but do not provide detailed sequence information. By contrast, long-read sequencing generates sequence information at the nucleotide level and offers unique advantages, including resolution of complex genomic regions, detection of structural variants at the base-pair level, and elucidation of epigenetic information^11–13^. While long-read sequencing has significantly improved our ability to analyze repeat expansions, several key questions regarding OPDM pathogenesis remain unresolved. Leveraging long-read sequencing technology, we aimed to investigate: (1) genotype/epigenotype-phenotype correlations, (2) sequence deviations within/flanking repeat expansions, (3) the role of asymptomatic carriers and their epigenetic features, (4) potential founder effects, and (5) mechanisms underlying repeat instability. To address these issues, we developed a novel sequencing approach using Nanopore CRISPR/Cas9-targeted resequencing (nCATS), which enables precise repeat length quantification, nucleotide variation detection, haplotyping, and CpG methylation assessment using genomic DNA samples.

## Results

### CGG repeat analysis

Nanopore CRISPR/Cas9-targeted resequencing was conducted using samples from a cohort comprising 71 individuals with OPDM, who are identified to have the expansion in *LRP12* (OPDM_LRP12; 7 families, 53 sporadic cases), 10 with OPDM_GIPC1 (2 families, 4 sporadic cases), and 8 with OPDM_NOTCH2NLC (8 sporadic cases) on Southern blotting. Pairs of guide RNAs were designed to flank the repeat regions in the four causative genes *(LRP12*, *GIPC1*, *NOTCH2NLC*, and *RILPL1*) (**Fig. 1A**). Mean read counts per patient was 19.8, with expansion accounting for 50.4% of total reads, consistent with heterozygous inheritance (**Supplementary Table 1**). Sequence data were collected from 1,008 expanded reads and 2,962 non-expanded reads. Expanded reads were detected in the same genes as those detected by Southern blotting (**Fig. 1B**). Almost all expanded reads contained uniform CGG repeats, and showed intra-patient size variability, particularly in samples from patients with OPDM_LRP12 and OPDM_NOTCH2NLC, indicating somatic instability of the repeats. Interestingly, we found that six patients had repeat expansions in two distinct genes, with three cases each having expansions in both *LRP12* and *GIPC1* or *LRP12* and *NOTCH2NLC* (**Fig.1B, Supplementary Fig. 1**).

**Fig. 1:**
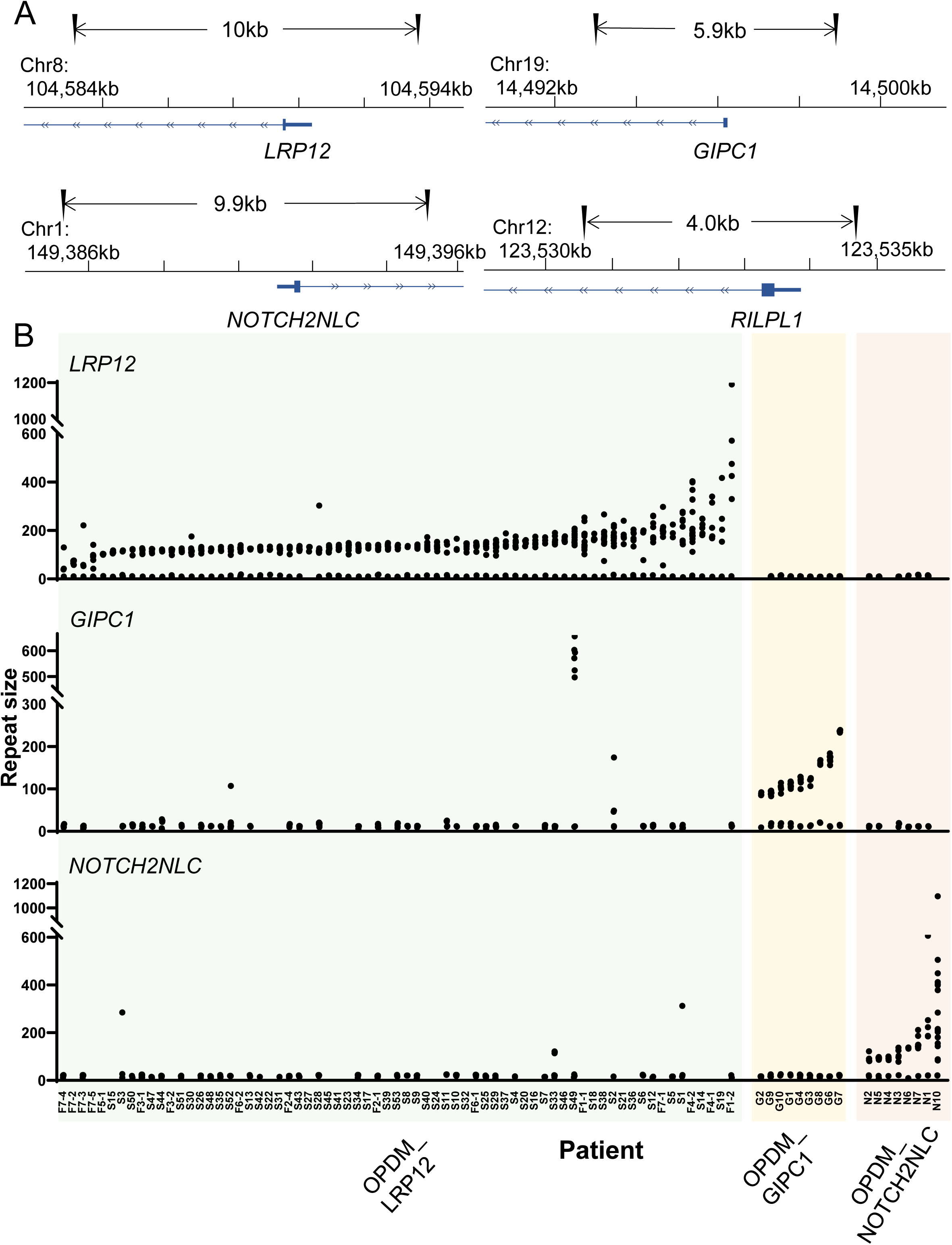
Nanopore CRISPR/Cas9-targeted resequencing design and distribution of repeat lengths in each causative gene in patients with oculopharyngodistal myopathy (OPDM). **A** Schematic illustrating the positions of pairs of guide RNAs designed to target sites flanking CGG repeats in each of four oculopharyngodistal myopathy causative genes: *LRP12*, *GIPC1*, *NOTCH2NLC*, and *RILPL1*. **B** Plots illustrating the repeat-size distributions in each causative gene in patients with OPDM. The horizontal-axis represents individual patients who diagnosed with *LRP12*, *GIPC1*, or *NOTCH2NLC* expansions on Southern blotting. Dots indicate observed repeat sizes (the nucleotide length in repeat region divided by three) of normal-sized reads alongside those of repeat-expanded reads. A wide range of lengths was observed, even within individual patients.

### Haplotype and flanking region analyses

Apparent geographical differences, with higher rates of expansions in *LRP12* or *GIPC1* detected in patients from eastern Asia^8^, led us to explore whether there was evidence of founder effects in OPDM. To assess the genetic backgrounds of patients with OPDM, we analyzed haplotypes associated with repeat expansions in *LRP12*, *GIPC1*, and *NOTCH2NLC* by constructing consensus allele sequences using expanded and non-expanded read data from each patient and comparing them with reference sequences. Among patients with OPDM_LRP12, we identified 10 single nucleotide variants (SNVs) that were present in 82.9% of expanded alleles but only 2.9% of non-expanded alleles (**Fig. 2A, Supplementary Fig. 2A**). Further, we identified seven SNVs present in repeat-expanded alleles in all patients (100%) with OPDM_GIPC1, which occurred in only 7.0% of non-expanded alleles (**Fig. 2B**, **Supplementary Fig. 2B**). Haplotypes including all 10 *LRP12* SNVs and 7 *GIPC1* SNVs were detected in 61 (2.63%) and 269 (11.58%), respectively, of 2,324 alleles in the general population, suggesting that the SNVs in each gene are in linkage disequilibrium in the Japanese population. Additionally, PacBio HiFi long-read sequencing data, generated from a subset of general population samples (n = 1162 individuals) showed that small repeat expansions ≥ 80 bp occurred only in haplotypes containing all 10 *LRP12* SNVs (**Supplementary Fig. 3A**) or 7 *GIPC1* SNVs (**Supplementary Fig. 3B**). Nevertheless, alleles with significantly expanded repeats were rare, suggesting a potential genetic predisposition to OPDM or a founder effect. No common SNV patterns were detected in expanded *NOTCH2NLC* alleles in patients with OPDM_NOTCH2NLC.

**Fig. 2:**
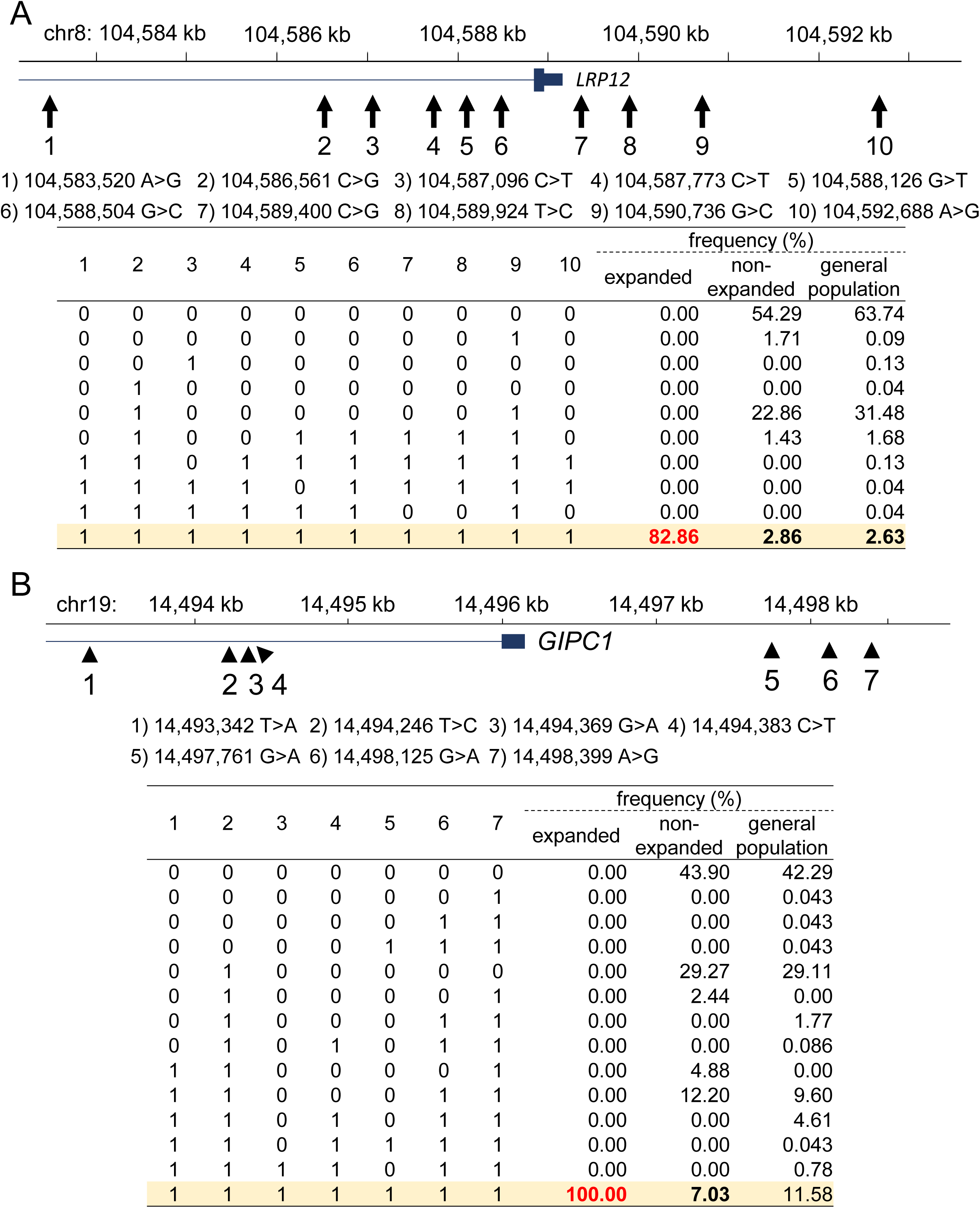
Haplotype analysis in patients with OPDM_LRP12. **A** Frequencies of specific haplotypes defined by 10 *LRP12* single nucleotide variants (SNVs) in expanded and unexpanded alleles in patients with OPDM_LRP12 and the general Japanese population. **B** Frequencies of specific haplotypes defined by 7 *GIPC1* SNVs in expanded and non-expanded alleles in patients with OPDM_GIPC1. **A**, **B** SNVs and their positions are described in top; SNV reference nucleotides are indicated by 0 and alternative nucleotides by 1 in the tables.

To further explore the differences between repeat-expanded and non-expanded alleles, we searched for differences in the sequences upstream and downstream of CGG repeat regions, and designated these as consensus and alternative flanking sequences shown in red and green, respectively in **Fig. 3**. In *LRP12*, non-expanded alleles harbored alternative flanking sequences at both sides of repeats, regardless of haplotype, whereas expanded alleles lacked these sequences, with the exception of two patients (S5 and S49) who only had the alternative flanking sequence in the region 5’ of the transcript (**Fig. 3A**). There were no obvious alternative flanking sequences in *GIPC1*, but expanded alleles contained a distinct repeat sequence in the 5’ region of the expanded transcript, which was consistently preserved within the same family (**Fig. 3B**). In *NOTCH2NLC*, no sequence differences specific to repeat-expanded or non-expanded alleles were detected (**Fig. 3C**). Structural prediction of single-strand RNA molecules in free minimum energy revealed that the identified flanking sequences significantly influenced the secondary structures of both *LRP12* and *GIPC1*, with notable differences between expanded and non-expanded alleles (**Supplementary Fig. 4A, B**). Interestingly, two patients with OPDM_LRP12 carried a non-expanded allele with the same haplotype as those of the repeat-expanded allele, while only expanded reads lacked the alternative flanking sequence (**Supplementary Fig. 5A, B**).

**Fig. 3:**
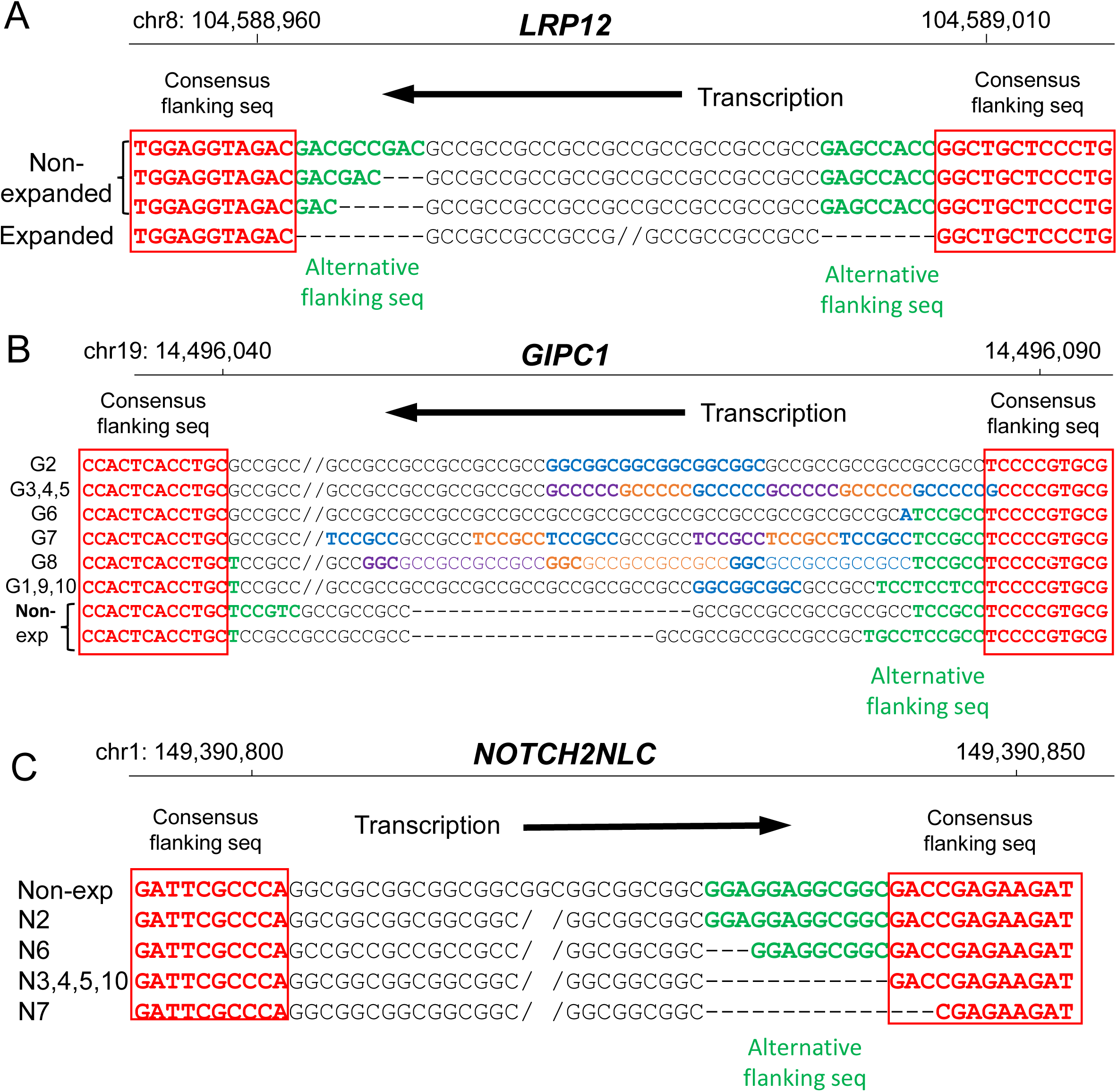
Flanking sequences around CGG repeats in three genes causative of oculopharyngodistal myopathy (OPDM). **A** *LRP12* flanking regions comprising consensus (red) and alternative (green) flanking sequences; non-expanded alleles contained one of the three alternative flanking sequences, whereas expanded alleles lacked them. **B** In *GIPC1*, repeat-expanded alleles harbored various distinct repeat sequences (in colored font) in their 5’-upstream regions, relative to transcription of the repeat. Consensus flanking sequences are shown in red. Alternative flanking sequences (green) did not vary between non-expanded and expanded reads. **C** *NOTCH2NLC* consensus (red) and alternative (green) flanking sequences. No specific sequences were identified in expanded or non-expanded reads.

### Structural variations related to complex RAN-translated products

Sequencing analysis also revealed structural variations outside the repeat expansion regions. Deletions of 44– and 58-bases in a region upstream of the repeat expansion were identified in transcripts from two cases with OPDM_LRP12 (S49 and F5-1, respectively) (**Supplementary Fig. 6A, B**). Predicted RAN translation indicated that these deletions, as well as lack of the alternative flanking sequence, may induce frameshifts, with different products translated from sites near to the altered nucleotides (**Fig. 4A**). For example, translation of the polyglycine-frame sequence will not occur in patients S5 and F5-1, suggesting that CGG repeat expansions do not necessarily result in generation of polyglycine tracts in OPDM_LRP12. In patient G7, who was diagnosed with OPDM_GIPC1, a nucleotide sequence duplication encompassing the region from the repeat expansion to upstream of the transcript was observed in all reads containing repeat expansions, and transcript analysis indicated that an upstream transcription start site (TSS) was used (**Supplementary Fig. 7A, B**). Consequently, proteins containing two separate amino acid polymers derived from each CGG repeat expansion were predicted to be produced (**Fig. 4B**). Further, patient N1 with OPDM_NOTCH2NLC carried a 38-bp deletion in the flanking region downstream of the CGG repeat (**Supplementary Fig. 8**). The major *NOTCH2NLC* transcript in skeletal muscle (NM_001364012.2) includes an upstream open reading frame (ORF), which encodes only polyglycine, as well as the *NOTCH2NLC* ORF; however, the downstream deletion likely results in a frameshift, leading to formation of a fusion protein containing both polyglycine and NOTCH2NLC (**Fig. 4C**).

**Fig. 4:**
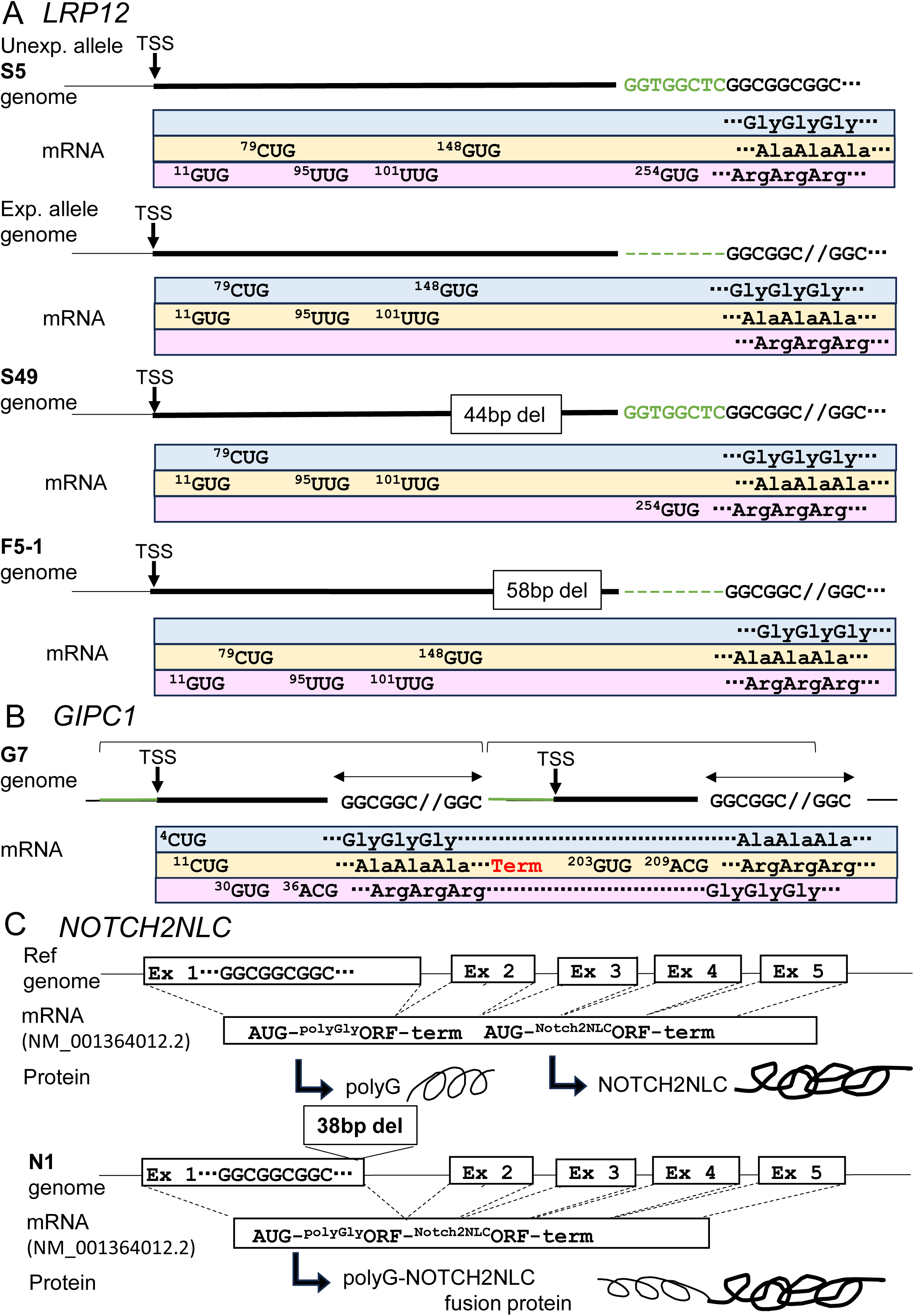
Predicted repeat-associated non-AUG (RAN) translated products from transcripts harboring structural variants. Schematics of genomic structure and near-cognate initiation sites in transcripts. **A** In *LRP12,* the non-expanded and expanded alleles had different frames of RAN translation. Polyglycine or polyalanine was translated from the CGG repeat region in an expanded transcript; however, in cases with deletions, frameshifts could occur, potentially preventing polyglycine production. **B** In cases with *GIPC1* duplication, an upstream TSS was used. Two tandem repeat expansions could produce three unique products, two containing different amino acid polymers. **C** The *NOTCH2NLC* skeletal muscle transcript (NM_001364012.2) contains two open reading frames that produce a glycine oligomer and NOTCH2NLC, respectively. An AUG codon upstream of the CGG repeat translates polyglycine from the repeat expansion. In cases with a 38-base deletion downstream of the repeat, the stop codon is lost, producing a fusion protein consisting of polyglycine and NOTCH2NLC.

### Genotype/epigenotype-phenotype correlations

To investigate intrafamilial repeat dynamics, we analyzed 12 individuals from four families with OPDM_LRP12 and three individuals from one family with OPDM_GIPC1 (**Supplementary Fig. 9**: this figure has been deleted from Supplemental materials in the preprint, because it can potentially make patients identifiable. The figure will be supplied on request), and detected a pattern of paternal contraction and maternal expansion of repeat length, suggesting genomic instability. Notably, two asymptomatic fathers of probands carried longer expanded repeats (ultra-long 459 ± 158 and 277 ± 101 repeats) beyond the range of those detected in symptomatic individuals (149 ± 37 repeats).

DNA methylation analysis demonstrated that asymptomatic carriers exhibited significantly higher levels of methylation in the ± 1 kb regions upstream and downstream of the TSS in ultra-expanded reads in OPDM_LRP12 and OPDM_GIPC1 families. In all patients, non-expanded reads were unmethylated, while expanded reads of pathogenic size in patients with OPDM_LRP12 were partially methylated, with substantial variation in methylation levels, and those from patients with OPDM_GIPC1 and OPDM_NOTCH2NLC exhibited relatively low levels of methylation (**Fig. 5A, Supplementary Table 2**). Further, we detected an intriguing pattern in the methylation rates of individual reads in samples from patients with OPDM_LRP12; at 1 kb upstream of the TSS, reads with ultra-long repeat sequences (> 300 repeats) were consistently hypermethylated, whereas the pathogenic expanded reads (< 300 repeats) exhibited a bimodal methylation pattern, where some were hypomethylated and others almost fully methylated (**Fig. 5B**). By contrast, samples from patients with OPDM_GIPC1 and OPDM_NOTCH2NLC exhibited low levels of methylation in pathogenic expanded reads (< 300 repeats for *GIPC1*, or < 600 repeats for *NOTCH2NLC*), although one patient (N7) with OPDM_NOTCH2NLC had exceptionally high methylation levels. These observations indicate gene-specific differences in rates of DNA methylation of the expanded regions, suggesting substantial differences in the epigenetic regulation of expanded alleles.

**Fig. 5:**
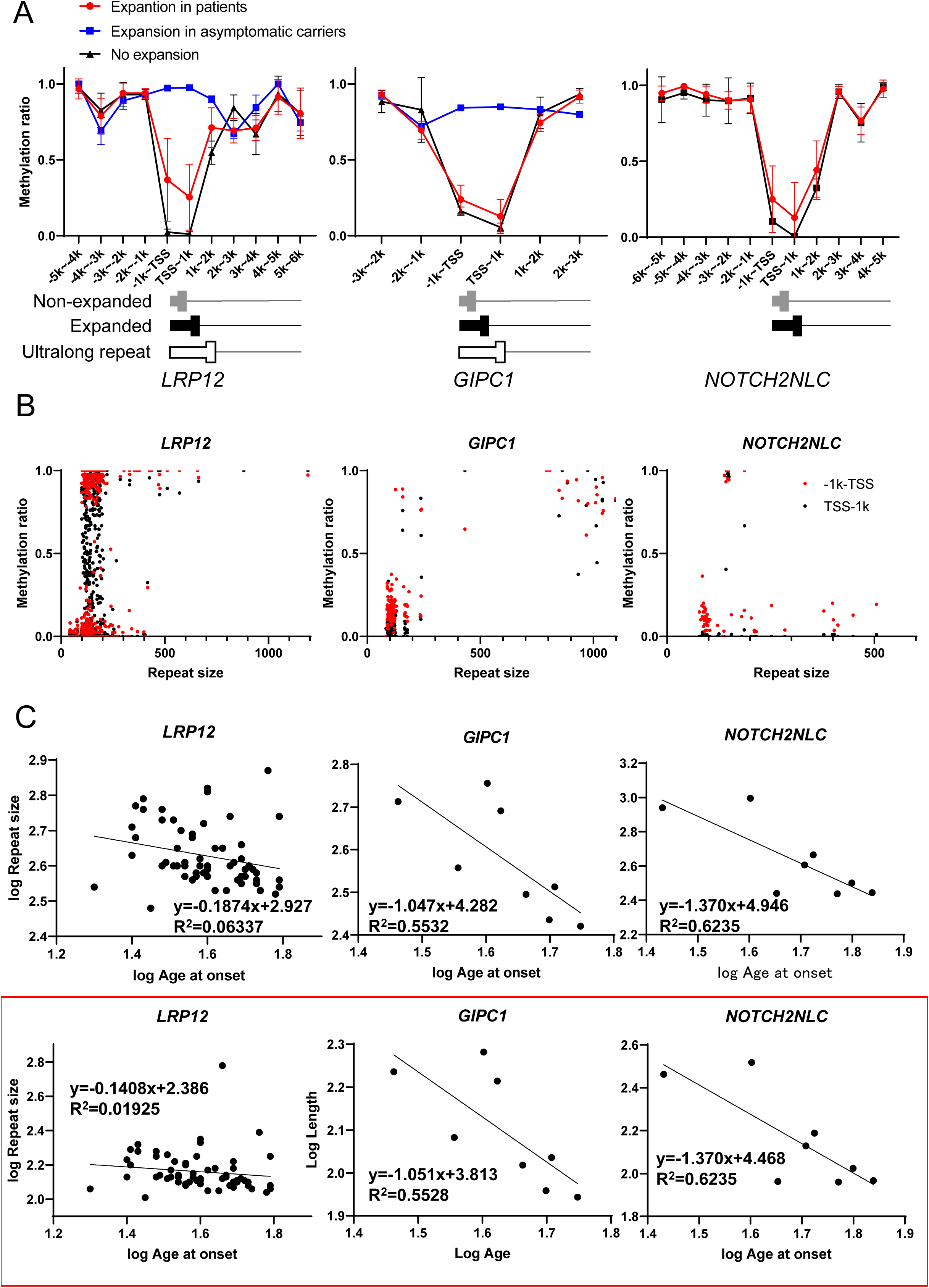
Genotype–phenotype correlations in oculopharyngodistal myopathy (OPDM). **A** Methylation rates in 1-kb sequence intervals based on the transcription start site (TSS) in non-expanded, expanded, and ultra-long expanded reads of each gene causative of OPDM. Methylation rates of non-expanded alleles in patients (black), expanded alleles in asymptomatic carriers (blue), and expanded alleles in patients (red). **B** Plots of CGG repeat size and methylation rates in each read across the three genes. In *LRP12*, DNA fragments with > 300 repeats were consistently hypermethylated, whereas those with < 300 repeats exhibited a bimodal methylation pattern, with some showing low methylation levels 1 kb upstream of the TSS, while others were highly methylated. Low methylation of almost all *GIPC1* or *NOTCH2NLC* fragments with pathogenic repeat sizes was observed. **C** Inverse correlations were observed between log (repeat length) and log (age of onset) values across all three genes.

Importantly, a significant inverted correlation between repeat length and age of onset was observed across all three genes, supporting the notion that longer repeat expansions are associated with earlier disease onset in patients with OPDM_GIPC1 or OPDM_NOTCH2NLC; however, the correlation coefficient was lower in patients with OPDM_LRP12, suggesting the presence of additional modifying factors (**Fig. 5C**).

As shown in Fig. 5A, the methylation levels in expanded reads in patients with OPDM_LRP12 were intermediate, with large error bars (*LRP12* in red); this highly variable CpG methylation rate in patients with OPDM_LRP12 may account for the lack of clear genotype– phenotype relationship in patients with this condition, relative to those in patients with OPDM_GIPC1 or OPDM_NOTCH2NLC. Interestingly, patients whose data plotted above the regression curve tended to have higher methylation rates (> 0.3), while those with data under the curve presented with lower rates (< 0.3) (**Fig. 6A**). Further, we found that methylation status varied from read-to-read within individual patients. **Fig. 6B** show the data that selected patients (S11, S9, F6-1, and S4) exhibited almost all-or-none patterns of expanded read methylation. These results suggest that total methylation rates of expanded reads in patients with OPDM_LRP12 may be determined by the proportions of fully methylated reads. This phenomenon could explain why the correlation between *LRP12* repeat length and age of onset was weaker, highlighting the complexity of genetic and epigenetic contributions to OPDM pathogenesis.

**Fig. 6:**
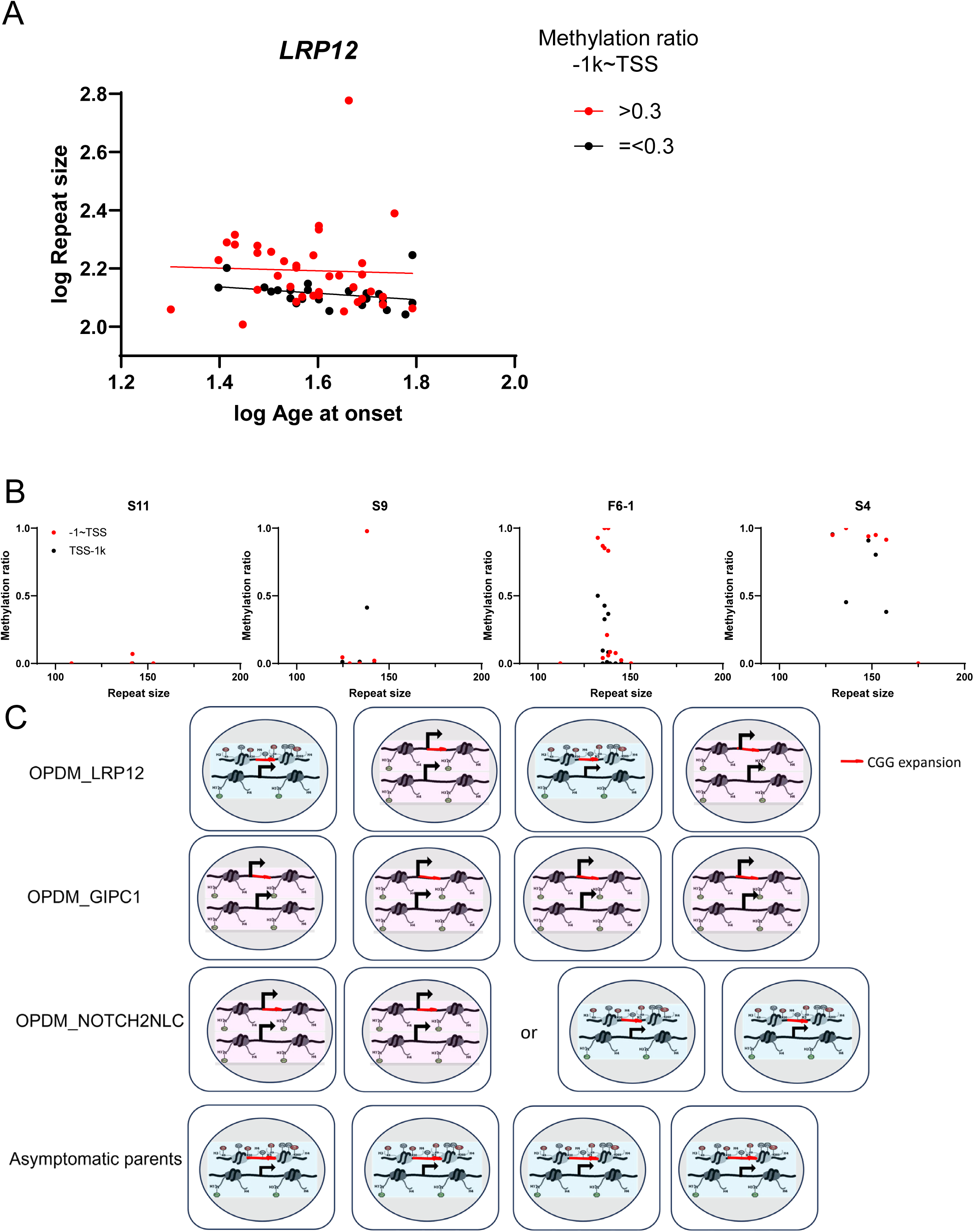
DNA methylation and genotype–phenotype correlations A Scatter plots showing the weaker correlation between log (CGG repeat size) and log (age of onset) values in patients with *LRP12* repeat expansions. Patients with mean methylation rates > 0.3 are shown in red, while those ≤ 0.3 are in black. Most red dots were plotted above the regression line, indicating higher methylation levels in reads with longer repeats. Square correlation coefficient and a formula for the linear regression were given by calculating of black dots showing lower significance. **B** Size and methylation rates around transcription start sites of individual expanded reads in samples from patients with OPDM_LRP12 (S11, S9, F6-1, and S4); plots show almost all-or-none methylation patterns in each patient. **C** Illustration of allele-specific methylation patterns of the patients across *LRP12*, *GIPC1*, *NOTCH2NLC*, and their asymptomatic parents. In *LRP12* patients, methylation levels varied among expanded alleles within the same individual, whereas *GIPC1* and *NOTCH2NLC* patients exhibited low methylation patterns across expanded alleles, except for one *NOTCH2NLC* patient. Blue cells represent high methylation in the expanded allele, likely associated with low expression rate. Red cells represent low methylation in the expanded allele, likely related to normal expression rate.

## Discussion

In this study, we adopted a novel approach to analyzing repeat expansions in patients with OPDM using single-molecule nCATS, enabling precise determination of repeat size, sequence variation, and methylation status. Our findings provide many new insights into OPDM pathogenesis, including genotype–phenotype relationships, founder effects, and potential mechanisms underlying repeat instability^1, 14^.

We found intra-patient repeat size variability, particularly in individuals with OPDM_LRP12. This variability is likely due to somatic instability of the expanded repeats, and may contribute to the clinical heterogeneity observed even within the same family. Additionally, we observed possible meiotic instabilities, such as a maternal bias toward repeat expansion and a paternal bias toward contraction, consistent with previous reports of intergenerational genomic instability^4, 10, 15^, and emphasizing the influence of parent-of-origin effects on repeat dynamics.

Repeat expansions in two genes were observed in six cases (**Supplementary Fig. 1**). In three cases (S52, S1, and S3), the proportions of expanded reads in the second gene among those harboring the same haplotype was extremely low, suggesting de novo expansion and a negligible contribution to disease pathogenesis. In another two cases (S2 and S33), the repeat expansion in the second gene was limited to 30 repeats, which does not reach the size range detected in patients and has also rarely been observed in the general population, indicating a lack of pathogenic significance^1^. In one case (S49), a repeat markedly expanded beyond the pathogenic size was detected in *GIPC1*, but all sequencing reads exhibited extensive methylation, suggesting that only the repeat expansion in *LRP12* contributed to disease pathogenesis (**Supplementary Table 3**).

Haplotype analysis identified a specific set of SNVs associated with repeat expansions in *LRP12* and *GIPC1*, suggesting genetic homogeneities with founder effects in patients and explaining the regional clustering of patients with OPDM in East Asia; however, we also found that these sets of 10 SNVs in *LRP12* and 7 SNVs in *GIPC1* are found at low frequencies in non-expanded reads and the general population, suggesting that presence of these haplotypes alone is not directly associated with repeat expansion. Moreover, we discovered that repeat-expanded reads lacked the flanking sequences found in non-expanded *LRP12* alleles, consistent with the findings of a recent study on *FGF14*, a gene implicated in spinocerebellar ataxia 27B, where flanking sequences differ between expanded and non-expanded alleles^16, 17^. These observations suggest that sequences flanking unexpanded alleles may contribute to maintenance of repeat length stability during meiosis, and that repeat expansion could occur in specific haplotypes, with repeat size extension accelerated by loss of flanking sequences. Indeed, two cases were identified that had the same pathogenic haplotype in repeat-expanded and non-expanded reads, but different flanking sequences, providing direct evidence that flanking sequences play a crucial role in repeat expansion. Although the precise mechanism underlying this phenomenon remains unclear, single-strand RNA structural predictions with/without flanking sequences revealed distinct secondary structures between expanded and non-expanded alleles.

Unlike *LRP12*, *GIPC1*, exhibited chimeric repeat sequences in the transcription direction 5′ region, which differed among patients but were preserved within two families across generations, despite differences in repeat length. Structural prediction further supported repeat instability, with the 3’-region lacking base-pairing potential. These findings expand our understanding of the molecular mechanisms driving repeat expansion and present new directions for further research^4, 9, 18^.

The CGG repeat expansion in the *NOTCH2NLC* skeletal muscle transcript is translated into a long glycine polymer, contributing to the protein aggregation underlying disease pathogenesis^19^. Unlike *NOTCH2NLC*, there is no ATG codon in *LRP12* upstream of the CGG repeats, making RAN translation a plausible alternative mechanism. Among near-cognate codons similar to the canonical AUG start codon, CUG is reported to be the most efficient for translation initiation, followed by GUG, ACG, and AUU^20^; therefore, transcripts with expanded repeats would theoretically only be expected to generate polyglycine or polyalanine from *LRP12* (**Fig4A,** Exp. allele). Interestingly, in patients with carrying a 58-base deletion upstream of the repeat (F5-1) or an alternative flanking sequence (S5) in *LRP12*, the glycine-frame would not be translated due to a frameshift. Similarly, in an OPDM_GIPC1 case (G7) with a duplication, an upstream TSS was used, leading to tandem translation of two expanded repeats and resulting in polyglycine-polyalanine, polyalanine, or polyarginine-polyglycine synthesis. The presence of these patients indicates that the RAN-translation theory is not a simple alternative to the polyglycine aggregation hypothesis as a universal mechanism for CGG repeat expansion disorders.

Although an inverted correlation was observed between repeat size and age of onset among patients with all three causative genes, the correlation was relatively weaker in patients with OPDM_LRP12, suggesting the presence of other modifying factors, including potential effects of epigenetic modification on gene expression. Interestingly, CpG methylation of the expanded repeat and flanking region was differentially regulated in each causative gene (**Fig. 6C**). Even within individual patients with expansion in *LRP12*, methylation levels differed in each expanded read, suggesting the methylation status differs in cell to cell. In contrast, in the patients with *GIPC1* or *NOTCH2NLC* expansion, almost all expanded reads remained unmethylated. Such differences in epigenetic modification increase the complexity of the genotype–phenotype relationship (**Fig. 5C**). This observation raises the possibility that the *LRP12* locus is intrinsically more susceptible to methylation upon repeat expansion. Interestingly, although the regression line between repeat length and age of onset among patients with non-methylated repeats (black dots, **Fig. 6A**) was shallower for *LRP12* than those for *GIPC1* and *NOTCH2NLC* **(Fig. 5C),** the range of the repeat length was also much shorter than that of the other causative genes. This finding suggests that even modest increases in repeat length exert a more pronounced phenotypic effect in *LRP12*, while longer expanded reads in *LRP12* undergo methylation, leading to transcriptional silencing, and resulting in attenuation of pathogenicity. The differential susceptibility to methylation among these genes may be attributable to variation in local chromatin context, including the presence or absence of regulatory elements, such as CTCF-binding sites, which influence DNA methylation and transcriptional insulation^18, 21^. Alternatively, it is possible that, because our cohort lacked data from individuals with disease onset at < 25 years-old, we failed to capture *LRP12* cases with long, unmethylated expansions. Collectively, these findings highlight the unique epigenetic landscape of *LRP12* and its role in shaping gene-specific genotype–phenotype correlations.

Our study had certain limitations. The nCATS analysis inherently generates fewer reads, which limits the ability to construct consensus sequences. Additionally, Nanopore long-read technologies are prone to sequencing errors; the error rate for CGG expansion in Nanopore sequencing in our study was 1.8% (data not shown) compared with 2.4% in a previous study^22^, highlighting the need for careful data interpretation.

Nevertheless, the nCATS technique provides precise information on the size, purity, and nucleotide patterns in triplet repeats, as well as on the structural variation and methylation state in/around repeats at the nucleotide level. Based on the data generated, we were able to explore complex genotype-epigenotype-clinical phenotype relationships, providing novel insights into the pathomechanism underlying OPDM diseases caused by short triplet repeat expansions, and paving the way for further investigations aimed at developing therapeutic strategies.

## Methods

### Subjects

Patients (n = 89) who were genetically diagnosed with OPDM using RP-PCR, AL-PCR, or Southern blotting, were selected from an in-house data repository and their clinical data reviewed, including: patients with an expansion in *LRP12* (OPDM_LRP12), n = 71; patients with an expansion in *GIPC1* (OPDM_GIPC1), n = 10; and patients with an expansion in *NOTCH2NLC* (OPDM_NOTCH2NLC), n = 8. This study was approved by the Ethical Committee of National Center of Neurology and Psychiatry, and all participants provided written informed consent.

### Genomic DNA preparation

Peripheral blood (10 ml) was mixed with 30 ml EL buffer (155 mM NH4Cl, 10 mM KHCO3, 1 mM EDTA, pH 7.4) on ice for 15 min, followed by centrifugation (KUBOTA 5930, RS-3012M) at 840 × g for 10 min at room temperature. After a repeat wash with EL buffer, pellets were suspended in 3 ml NL buffer (10 mM Tris–HCl, 2 mM EDTA, 400 mM NaCl, pH 8.2). Subsequently, 1% SDS and 20mg/ml proteinase K were added, and the mixture incubated at 37°C overnight. DNA lysis solution was then supplemented with 1 ml 5 M NaCl, followed by phenol/chloroform extraction and ethanol precipitation, and DNA pellets dissolved in TE buffer.

### DNA library preparation for nCATS

DNA libraries were prepared using a ligation sequencing kit (Oxford Nanopore Technologies, SQK-LSK109). Cas9 ribonucleoprotein complexes (RNPs) were generated by incubating a pool of annealed 1 μM tracrRNA-crRNA and 0.5 μM HiFi Cas9 (IDT, #1081061) at room temperature for 30 min. Pairs of guide RNAs targeting both sides of the repeat sites in the four causative genes (*LRP12*, *GIPC1*, *NOTCH2NLC,* and *RILPL1*), were designed using the CRISPR Custom Design Tool provided by Integrated DNA Technologies (IDT) (https://sg.idtdna.com/site/order/designtool/index/CRISPR_CUSTOM). Guide RNAs were synthesized by IDT (**Fig. 1A**, **Supplementary Table 4**). Genomic DNA (2 μg) was dephosphorylated with Quick Calf Intestinal Phosphatase (NEB, #M0525S) at 37°C for 10 min, followed by inactivation at 80°C for 2 min. The dephosphorylated genomic DNA was then treated with a mixture of Cas9 RNPs, Taq polymerase (NEB, #M0273S), and dATP (NEB, #N0440S) at 37°C for 30 min, followed by 72°C for 5 min for Cas9 RNP cleavage and dA-tailing. For native barcode ligation, a native barcoding expansion 1–12 kit (Oxford Nanopore Technologies, EXP-NBD104) was used to ligate barcodes to the cleaved and dA-tailed genomic DNA samples with Blunt/TA Ligase Master Mix (NEB, #M0367L) at room temperature for 10 min, followed by purification on a magnet using Agencourt AMPure XP Beads (Beckman Coulter, #A63880). AMII adapters were subsequently ligated to the barcoded genomic DNA using Quick T4 DNA ligase (NEB, #E6056S) at room temperature for 10 min, followed by purification on a magnet with AMPure XP Beads. The prepared DNA library was then loaded into a MinION Flow Cell (FLOMIN106D) on a MinION Mk1C, and sequencing performed for 20–21 h.

## Data analysis

Bases were called from Fast5 files using Guppy to generate Fastq files. Reads were aligned to the reference sequence (GRCh38) using Minimap2 and then visualized using Integrative Genomics Viewer^23, 24^. The sequences of repeats and their flanking regions were aligned to deduce consensus sequences using Genetyx software. Nanopolish was used for DNA methylation analysis^25^. The estimated number of methylated cytosines was calculated by multiplying the methylation rate obtained by Nanopolish by the number of CpGs. Genotype–phenotype correlations were assessed by comparing molecular data with clinical parameters, such as disease onset.

## Statistics

All statistical analyses were conducted using R (version 4.2.0) and GraphPad Prism (version 9.0). Correlations between repeat length and clinical parameters were calculated using Pearson’s correlation coefficient. Differences between groups were assessed by Student’s t-test or Mann-Whitney U test for two-group comparisons, and one-way ANOVA for multiple-group comparisons; p-values < 0.05 were considered statistically significant.

## List of abbreviations

OPDM, oculopharyngodistal myopathy; nCATS, Nanopore CRISPR/Cas9-targeted resequencing; RAN, repeat-associated non-AUG; RP-PCR, repeat-primed PCR; AL-PCR, amplicon-length analysis PCR; SNV, single nucleotide variant; TSS, transcription start site; ORF, open reading frame; RNPs, ribonucleoprotein complexes; IDT, Integrated DNA Technologies

## Declarations

### Ethics approval and consent to participate

All patients provided informed consent for use of their samples for research after diagnosis. This study was approved by the ethical committees of the NCNP (A2022-045).

## Availability of data and materials

The data supporting the findings in this study are available from the corresponding author upon request.

## Authors’ contributions

Conceptualization, SN; Formal analysis, NE, SN, MO, AY, KyS, and SH; Investigation, NE, SN, MO, and KyS; Methodology, NE and SN; Patient evaluations, collecting patient samples, and/or clinical data, NE, AY, KyS, SH, YO, and IN; Resources, YO and IN; Supervision, SN; Project administration, KaS and IN; Writing – original draft, NE and SN; Writing – reviewed, all authors.

## Data Availability

All data produced in the present study are available upon reasonable request to the authors

## Supplementary Information

**Supplementary Fig. 1: Data from patients with repeat expansions in two genes.**

**Supplementary Fig. 2: Specific haplotypes found in *LRP12* and *GIPC1* alleles with expanded reads.**

**Supplementary Fig. 3: Distribution of CGG repeat lengths in individuals with specific *LRP12* and *GIPC1* haplotypes in the general population in Japan.**

**Supplementary Fig. 4: RNA secondary structures of *LRP12* and *GIPC1* expansions and their flanking sequences.**

**Supplementary Fig. 5: Data from a patient with the same haplotype in both expanded and non-expanded *LRP12* alleles.**

**Supplementary Fig. 6: Structural variations in *LRP12* expanded reads in two patients.**

**Supplementary Fig. 7: Structural variation in *GIPC1* expanded reads and the transcription start site in patient G9.**

**Supplementary Fig. 8: Structural variation in the region downstream of the repeat expansion in *NOTCH2NLC*.**

**Supplementary Fig. 9: Intrafamilial dynamics of repeats in *LRP12* and *GIPC1*.**

**Supplementary Table 1. Clinical information, average size of expanded repeats, and numbers of obtained expanded and non-expanded reads obtained for each patient.**

**Supplementary Table 2. Average methylation rates in 1kb sequence windows around transcription start sites in expanded and non-expanded reads.**

**Supplementary Table 3. Average methylation rates in the expanded reads observed in the additional genes in six patients having repeat expansions in two distinct genes.**

**Supplementary Table 4. Guide RNA sequences for analysis of four causative genes by nCATS.**

**Supplementary Fig. 1:**
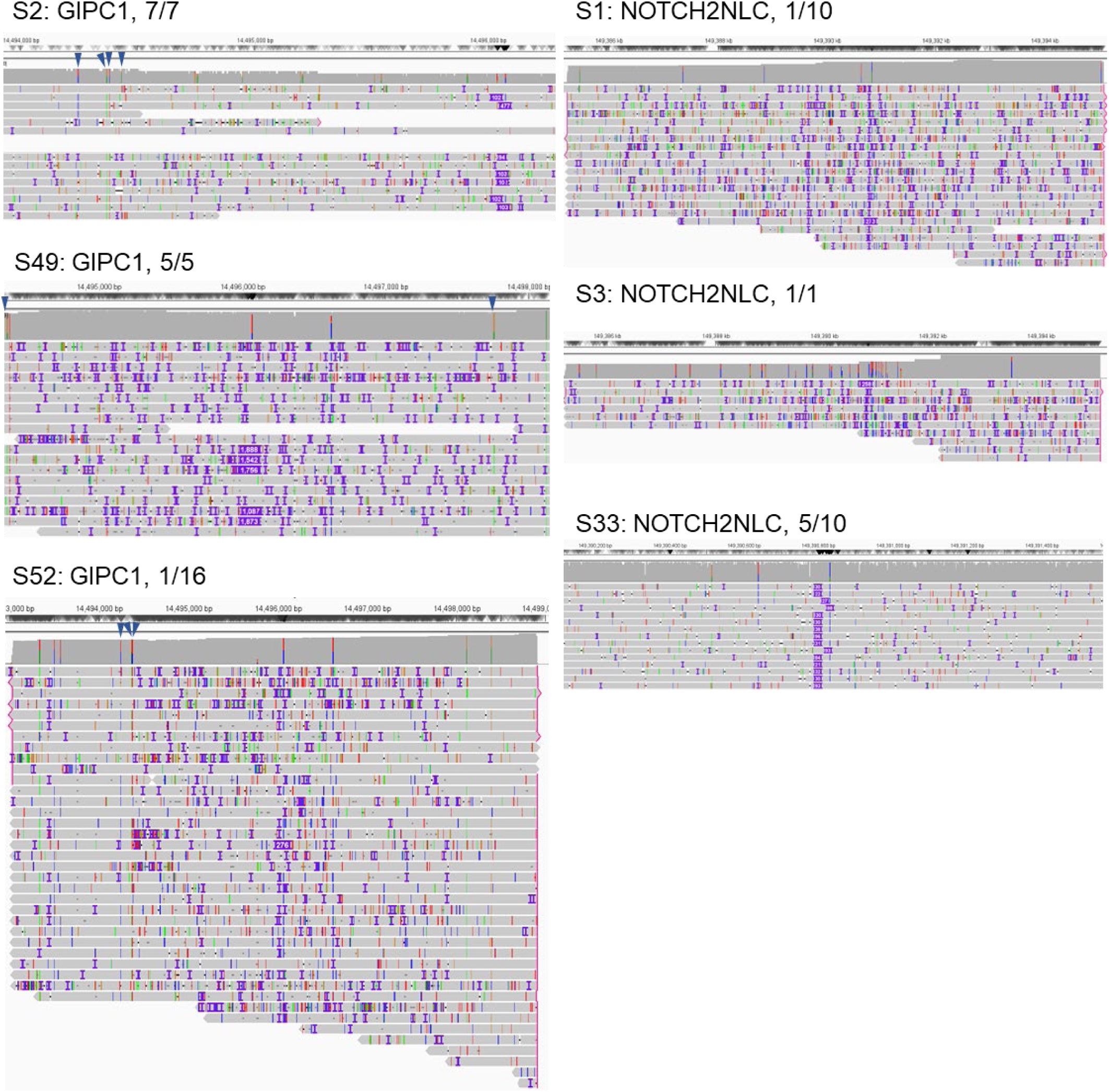
Data from patients with repeat expansions in two genes. Six patients had repeat expansions in two distinct genes. Panels show reads from genes other than the predicted causative genes. Three patients (S2, S49, and S52) had reads with repeat expansions in *GIPC1* in addition to causative expansions in *LRP12*, and three (S1, S3, and S33) had reads with repeat expansions in *NOTCH2NLC* in addition to causative expansions in *LRP12*. Numbers indicate the detection rates of reads derived from each allele.

**Supplementary Fig. 2:**
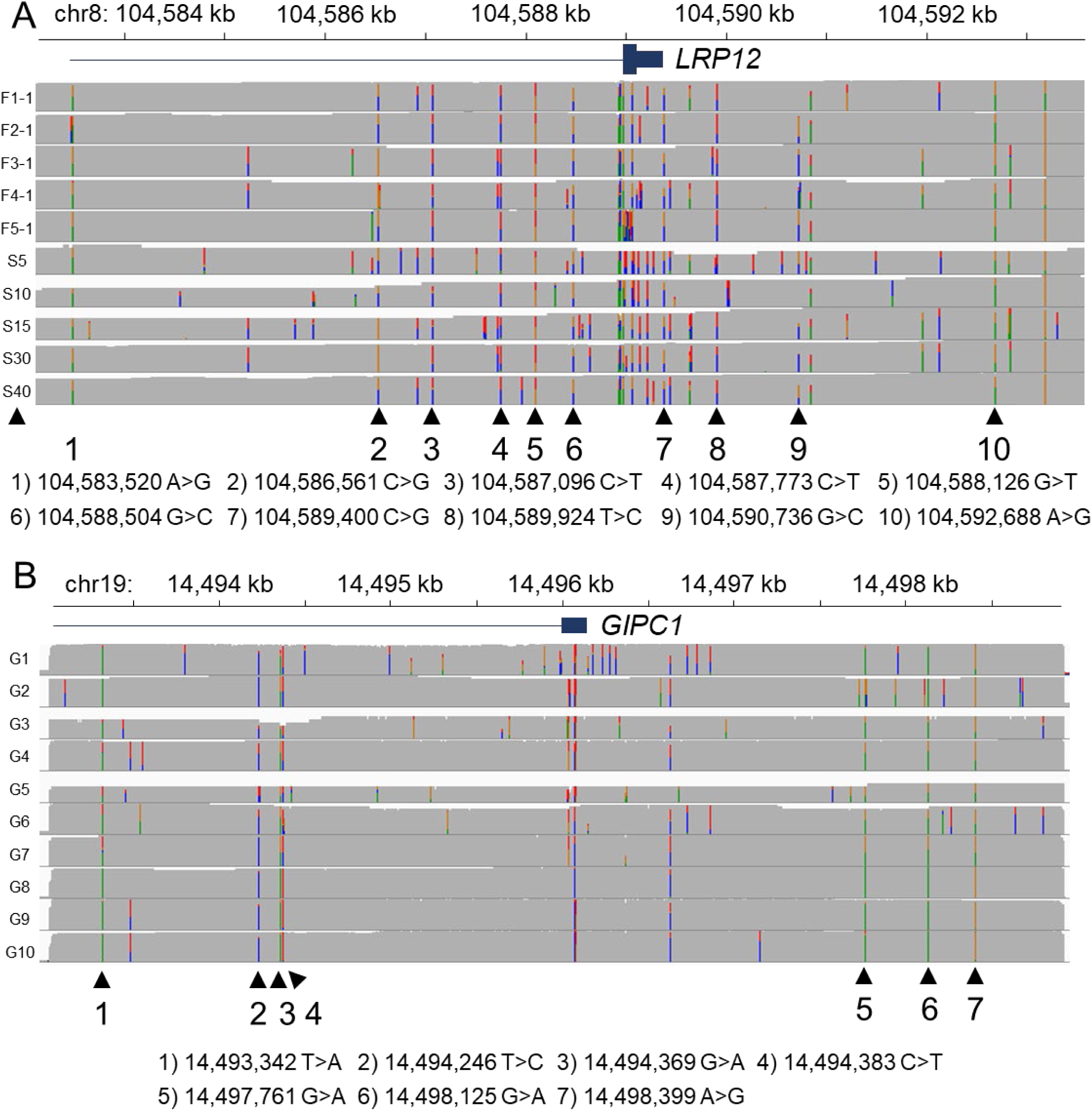
Specific haplotypes found in *LRP12* and *GIPC1* alleles with expanded reads. **A** Ten common single nucleotide variants (SNVs) detected in reads with *LRP12* repeat expansions; data from 11 patients are presented. **B** Seven common SNVs found in reads with *GIPC1* repeat expansions; data from 10 patients are presented.

**Supplementary Fig. 3:**
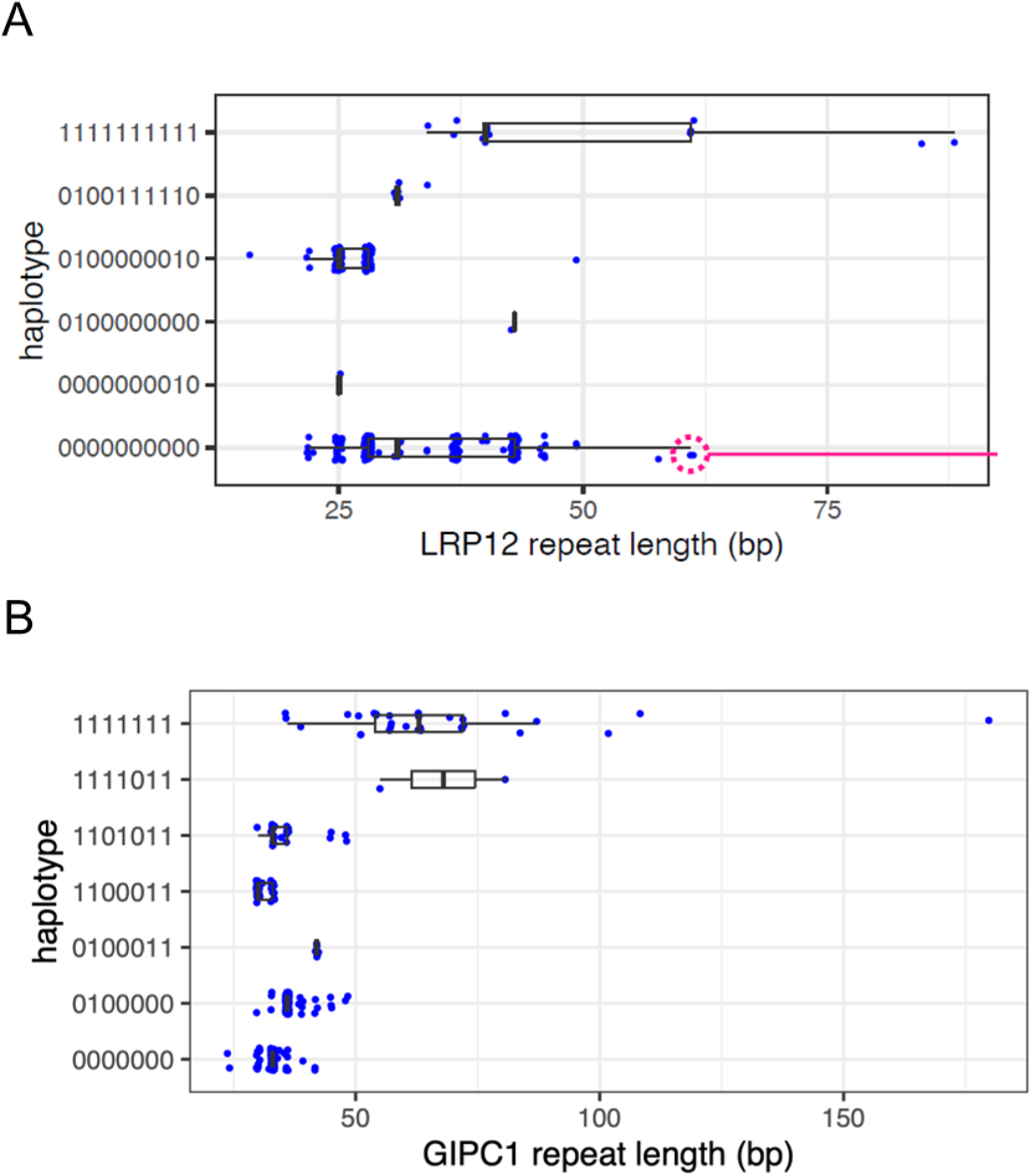
Distribution of CGG repeat lengths in individuals with specific *LRP12* and *GIPC1* haplotypes in the general population in Japan. (A) Distribution of CGG repeat lengths in different haplotypes defined by 10 SNV sites in *LRP12* in the general population. The specific haplotype with the 10 SNVs (all 1) had larger size of repeat length. (B) Distribution of CGG repeat lengths in haplotypes defined by 7 SNV sites in *GIPC1* in the general population. The specific haplotype with the 7 SNVs (all 1) had larger size of repeat length.

**Supplementary Fig. 4:**
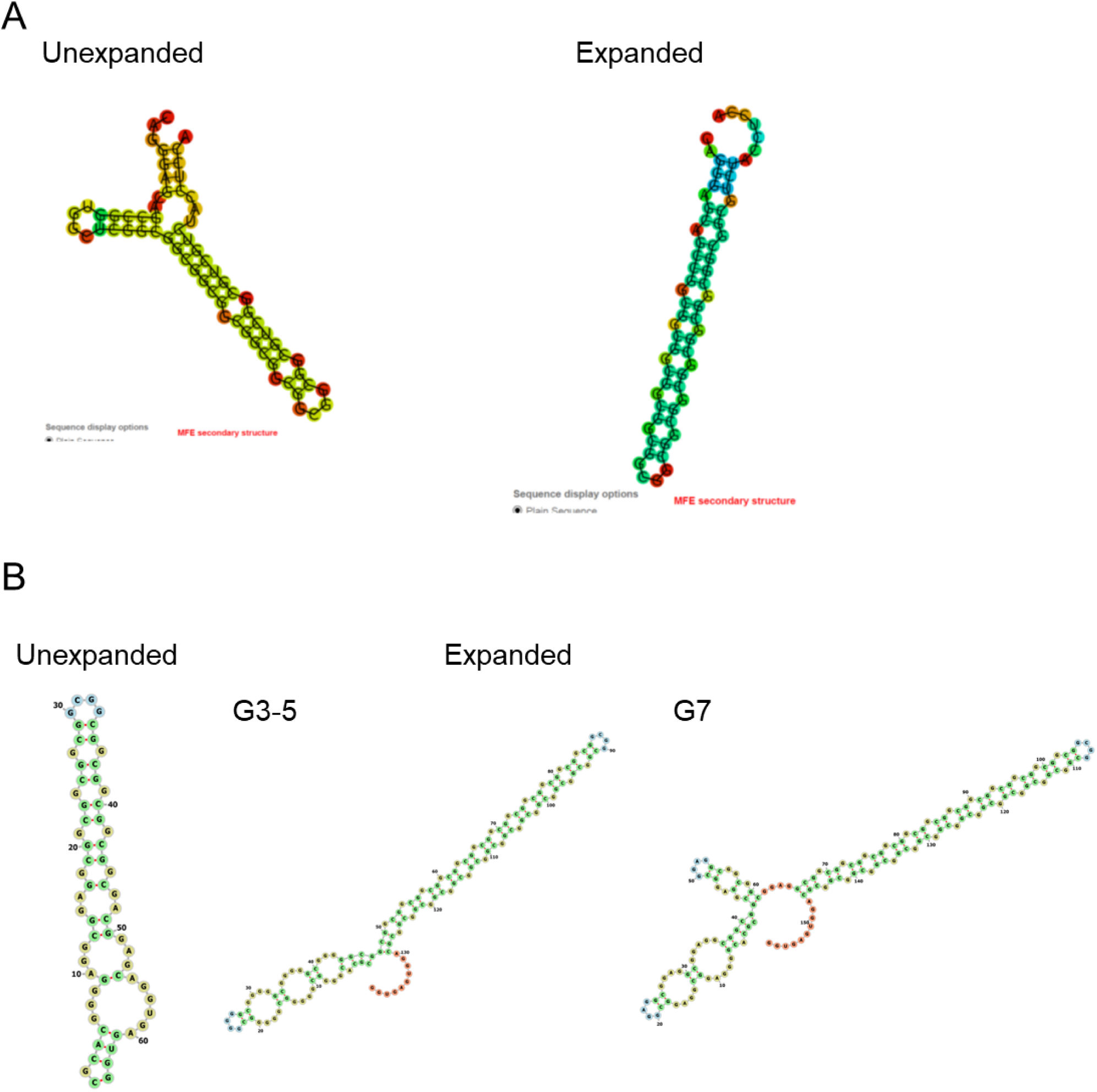
RNA secondary structures of *LRP12* and *GIPC1* expansions and their flanking sequences. **A** Predicted secondary structures of flanking sequences in expanded and non-expanded *LRP12* alleles. **B** Predicted secondary structures of flanking sequences in expanded and non-expanded *GIPC1* alleles.

**Supplementary Fig. 5:**
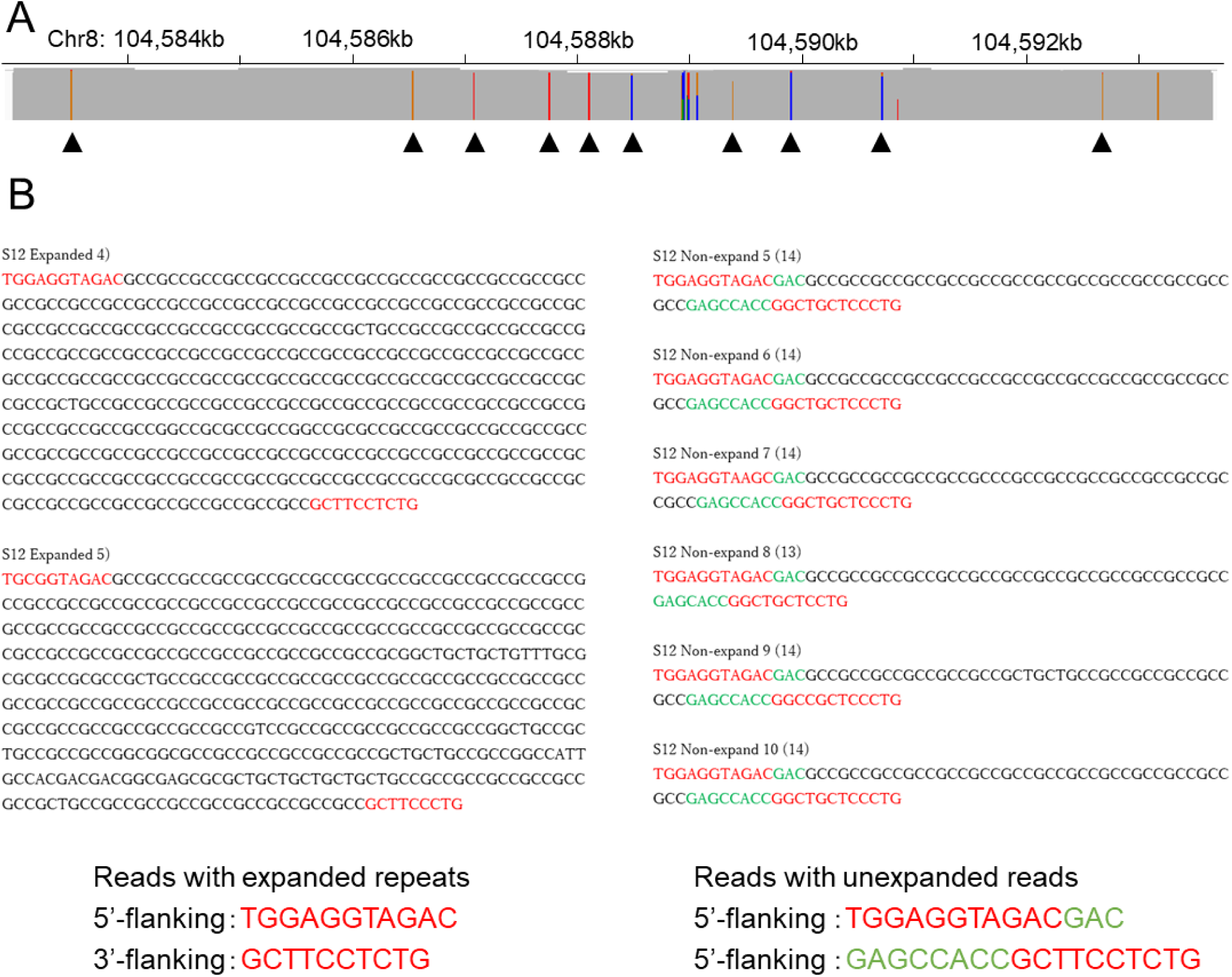
Data from a patient with the same haplotype in both expanded and non-expanded *LRP12* alleles. **A** *LRP12* single nucleotide variant (SNV) data from a patient with an *LRP12* expansion who was homozygous for all 10 *LRP12* SNVs (see Supplementary Fig. 1). **B** The expanded allele sequence showing lack of the flanking sequences (green), which were present in the non-expanded allele.

**Supplementary Fig. 6:**
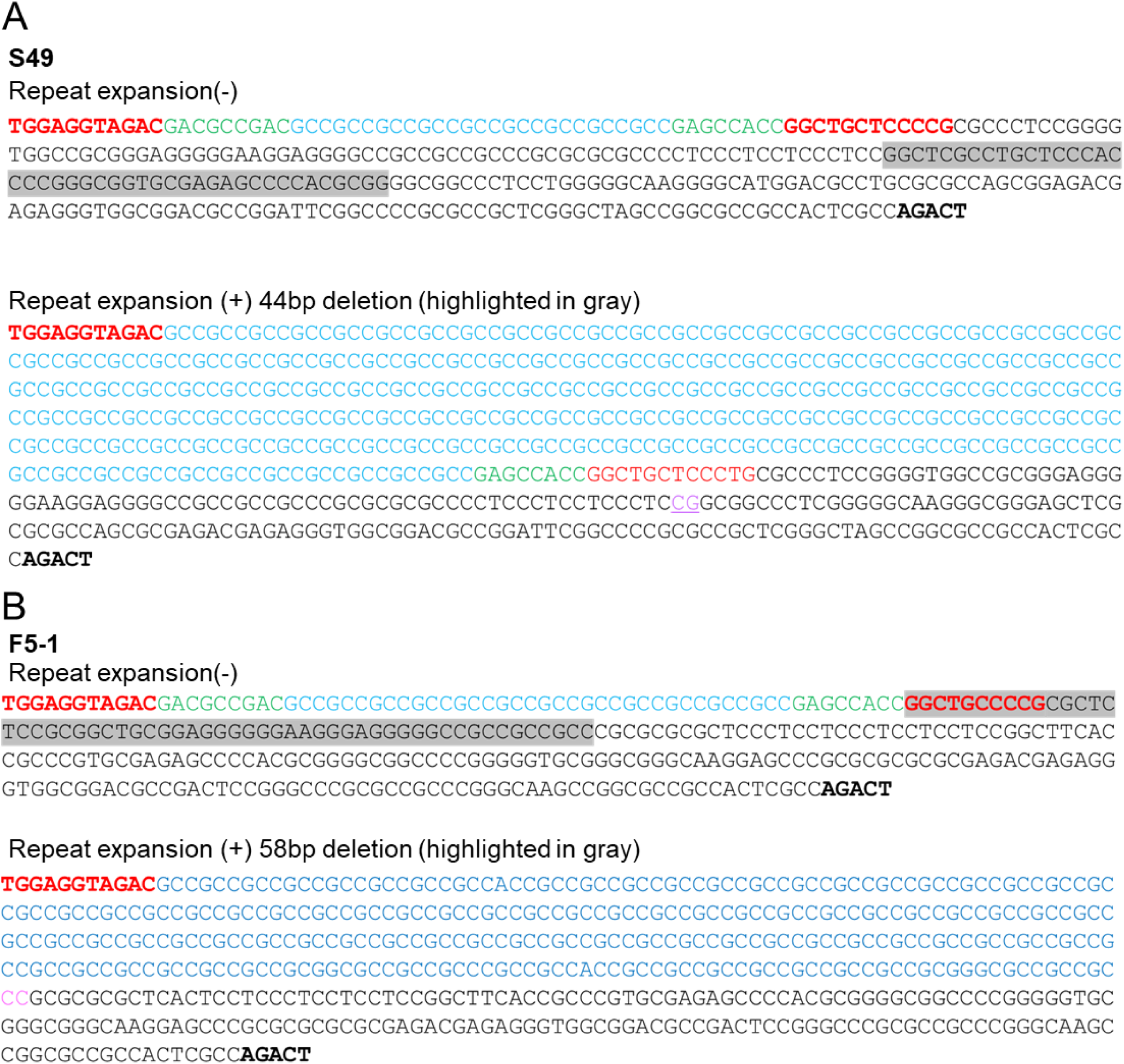
Structural variations in *LRP12* expanded reads in two patients. **A** A 44-bp deletion (highlighted gray in non-expanded reads) between the dinucleotides in pink upstream of the expansion (light blue) and flanking sequences (green and red) in the transcript direction in Patient S49. **B** A 58-bp deletion (highlighted gray in non-expanded reads) between the dinucleotides in pink upstream of the expansion (light blue) and flanking sequences (green and red) in the transcript direction in Patient F5-1. The transcription start site is indicated in bold.

**Supplementary Fig. 7:**
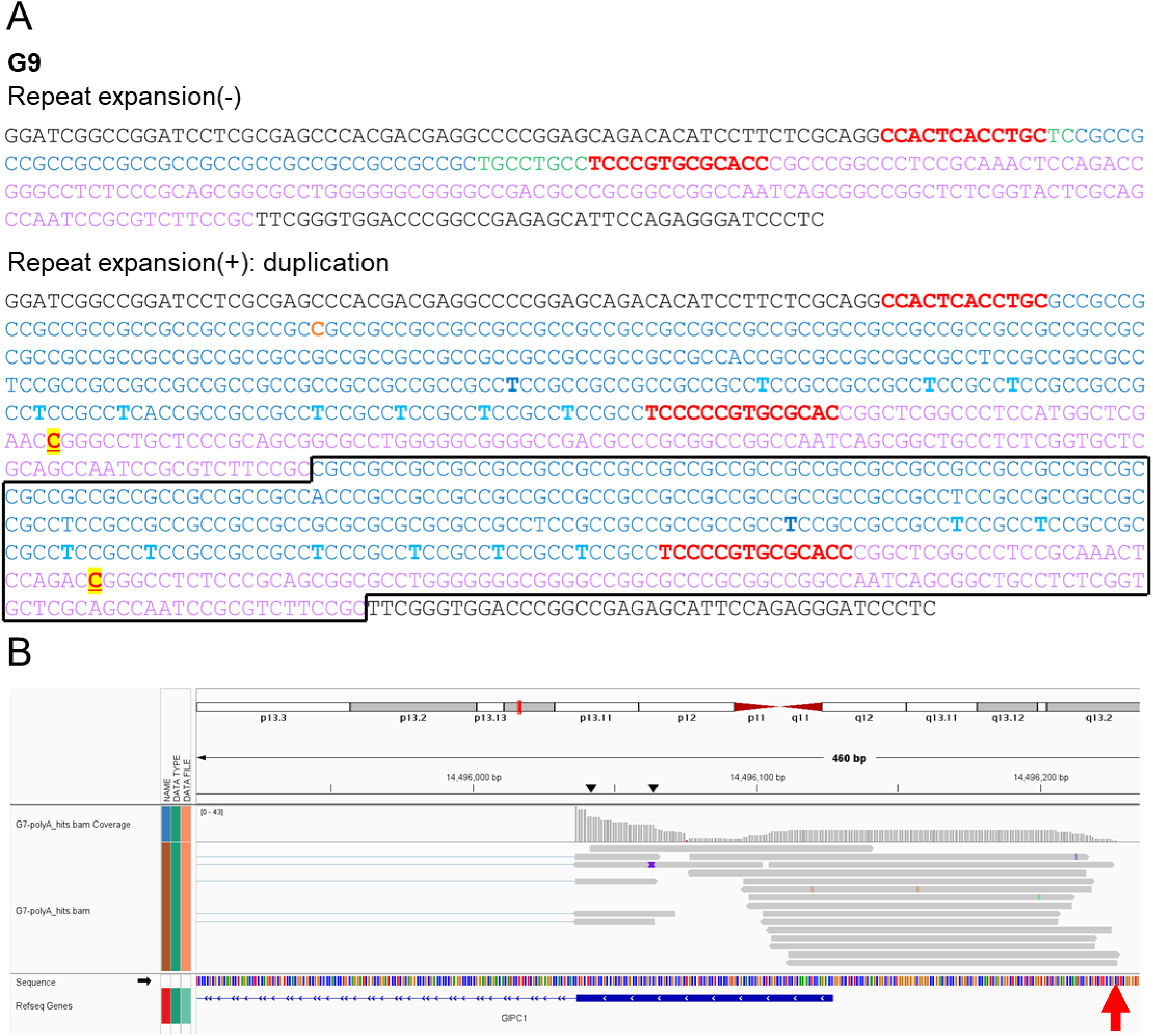
Structural variation in *GIPC1* expanded reads and the transcription start site in patient G7. **A** Sequence from patient G7, showing a duplication (black rectangle) including the expanded repeat sequence (light blue) and its 5’-upstream region (pink) in expanded *GIPC1* reads. **B** Transcript analysis of data from patient G7 revealing transcription initiation from an upstream transcriptional start site (arrow) located within the duplicated region.

**Supplementary Fig. 8:**
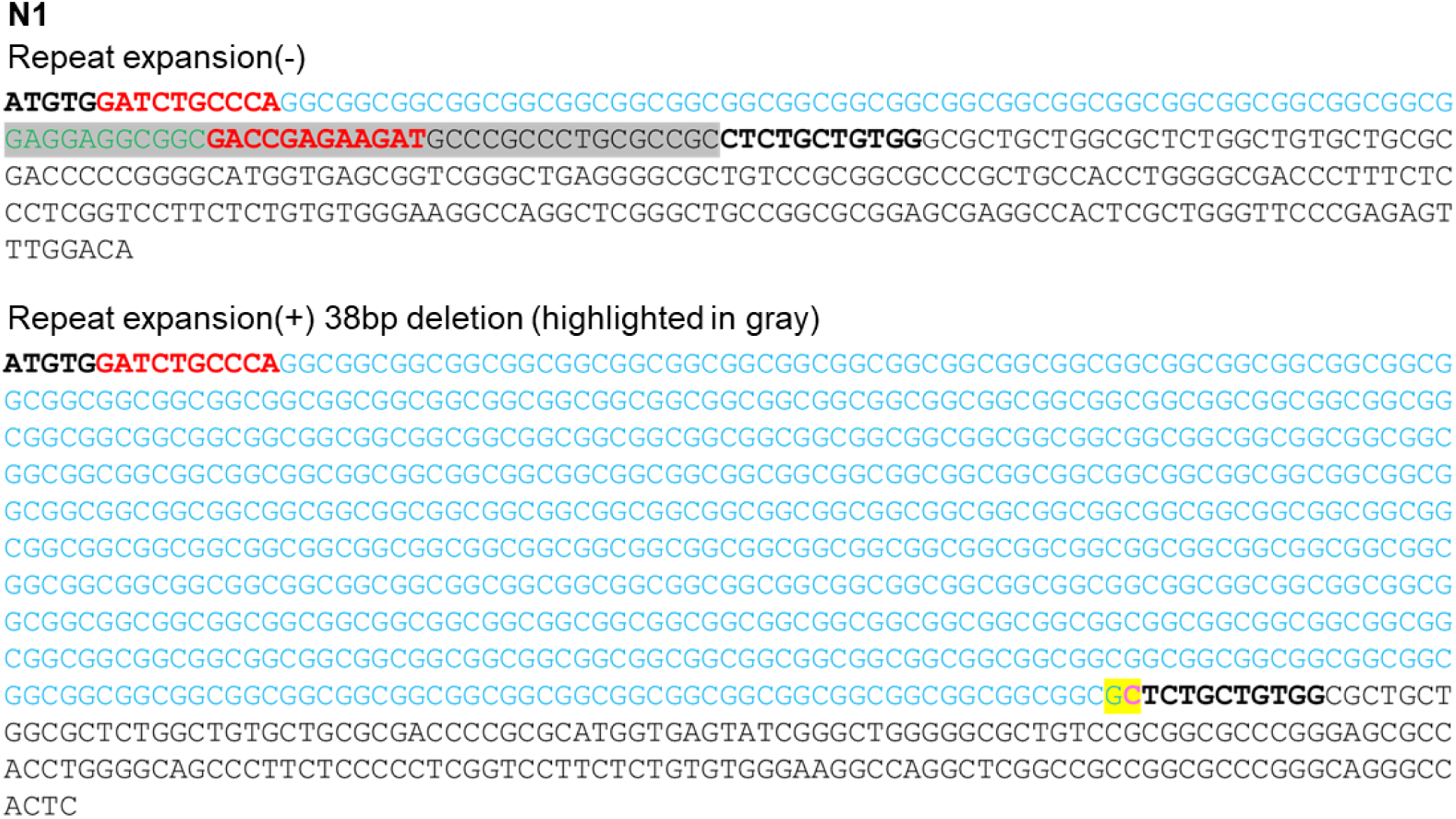
Structural variation in the region downstream of the repeat expansion in *NOTCH2NLC*. A patient with a 38-bp deletion (highlighted gray in non-repeat sequence) between the dinucleotides highlighted yellow downstream of the CGG repeats in *NOTCH2NLC*.

**Supplementary Table 1.**
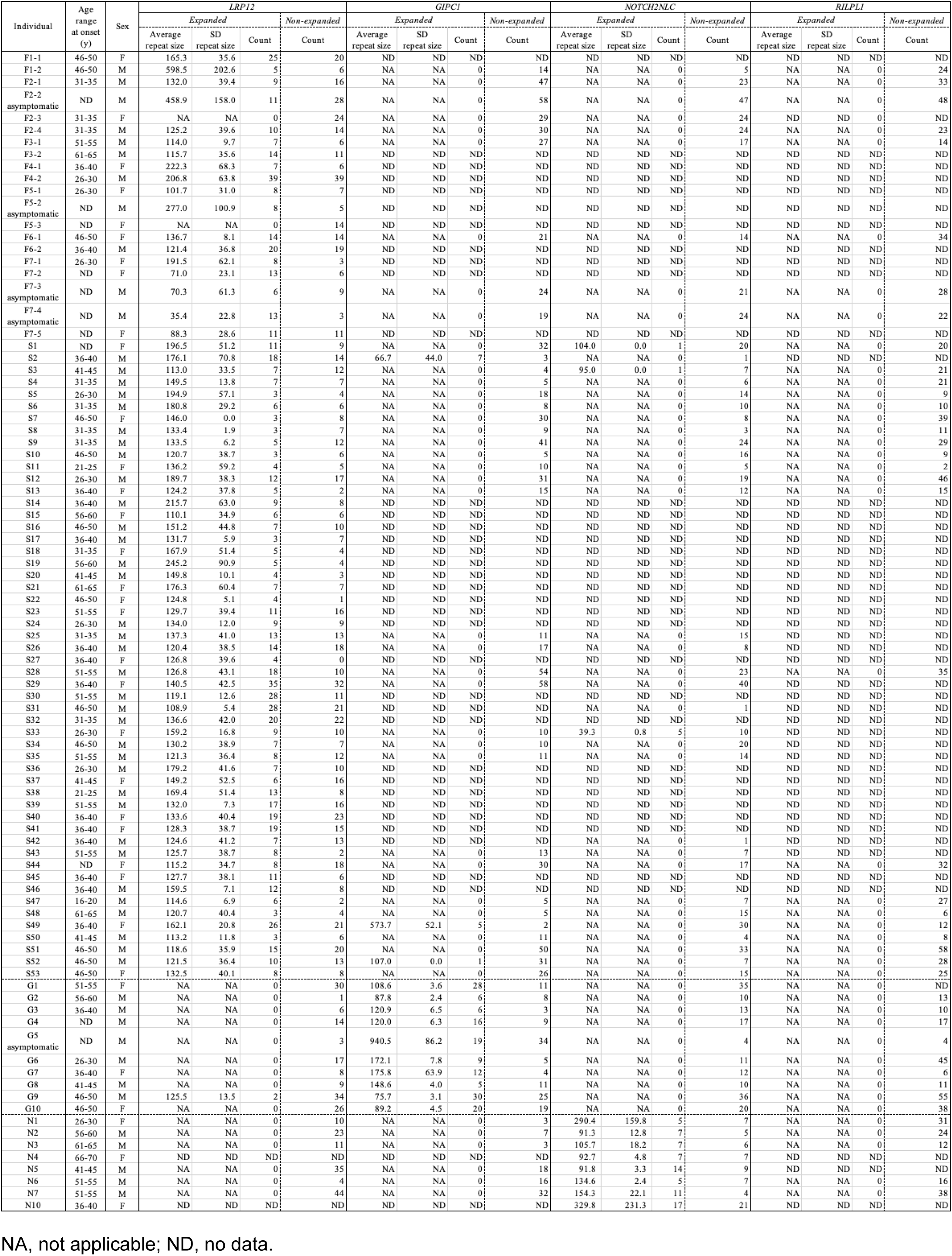
Clinical information, average size of expanded repeats, and numbers of obtained expanded and non-expanded reads obtained for each patient.

**Supplementary Table 2.**
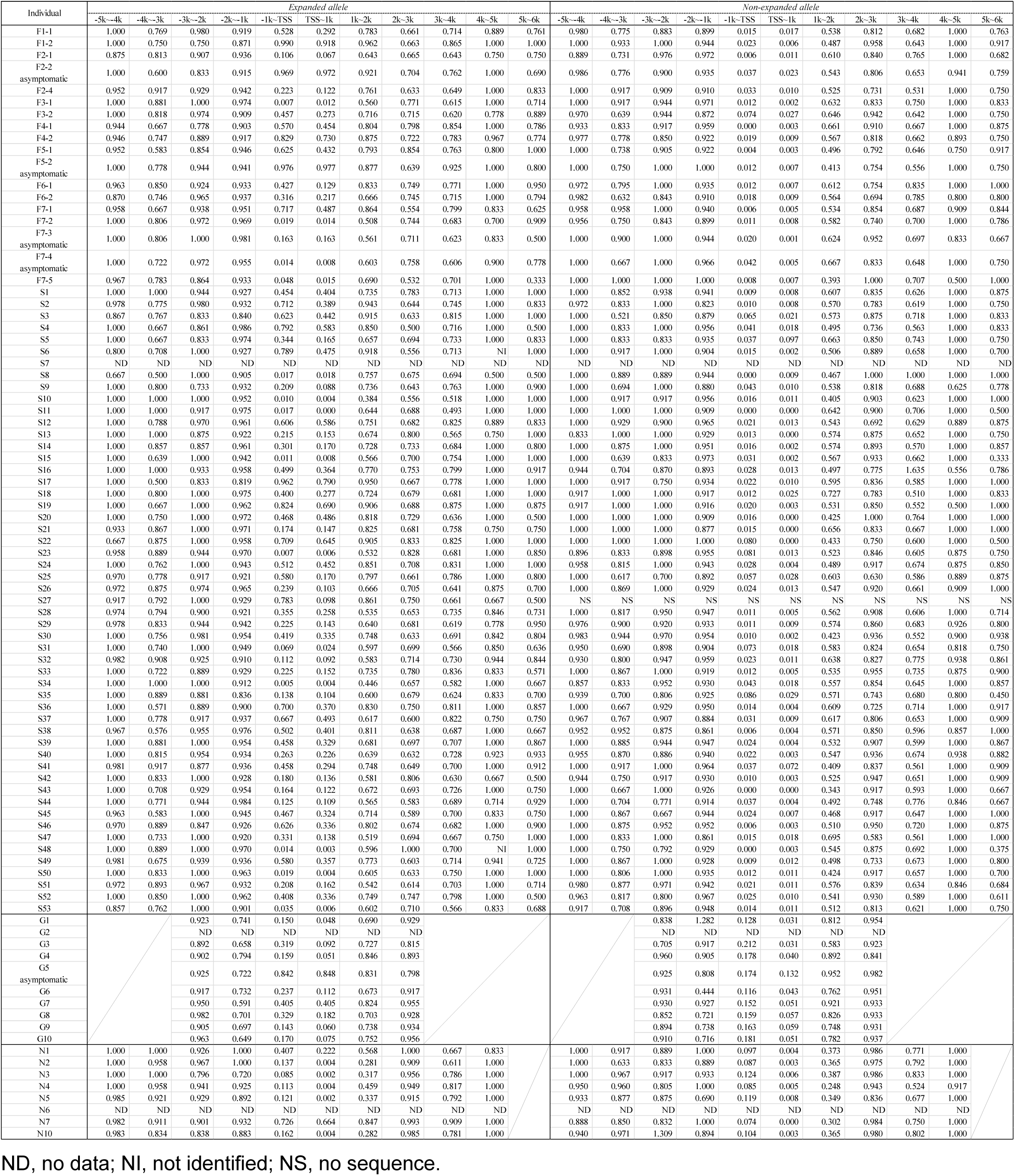
Average methylation rates in 1kb sequence windows around transcription start sites in expanded and non-expanded reads.

**Supplementary Table 3.** Average methylation rates in the expanded reads observed in the additional genes in six patients having repeat expansions in two distinct genes.

| Individual | Causative gene | Expanded allele |  |  |  |  |  |  |  |  |  |
| --- | --- | --- | --- | --- | --- | --- | --- | --- | --- | --- | --- |
|  |  | -5k~-4k | -4k~-3k | -3k~-2k | -2K~-1K | -1k~TSS | TSS~1k | 1k~2k | 2k~3k | 3k~4k | 4k~5k |
| S02 | GIPC1 |  |  | NI | 1.000 | 0.103 | 0.000 | 0.500 | 1.000 |  |  |
| S49 |  |  |  | 0.928 | 0.768 | 0.854 | 0.717 | 0.820 | 0.931 |  |  |
| S52 |  |  |  | 0.750 | 0.375 | 0.250 | 0.037 | 1.000 | 1.000 |  |  |
| S01 | NOTCH2NLC | NI | NI | 0.000 | 1.000 | 1.000 | 0.088 | 0.011 | 0.462 | 1.000 | 1.000 |
| S03 |  | 1.000 | 0.667 | 1.000 | 1.000 | 1.000 | 0.115 | 0.048 | 0.400 | 1.000 | 1.000 |
| S33 |  | 0.938 | 0.893 | 0.852 | 0.820 | 0.081 | 0.004 | 0.345 | 0.980 | 0.583 | 0.625 |

NI, not identified.

**Supplementary Table 4.**
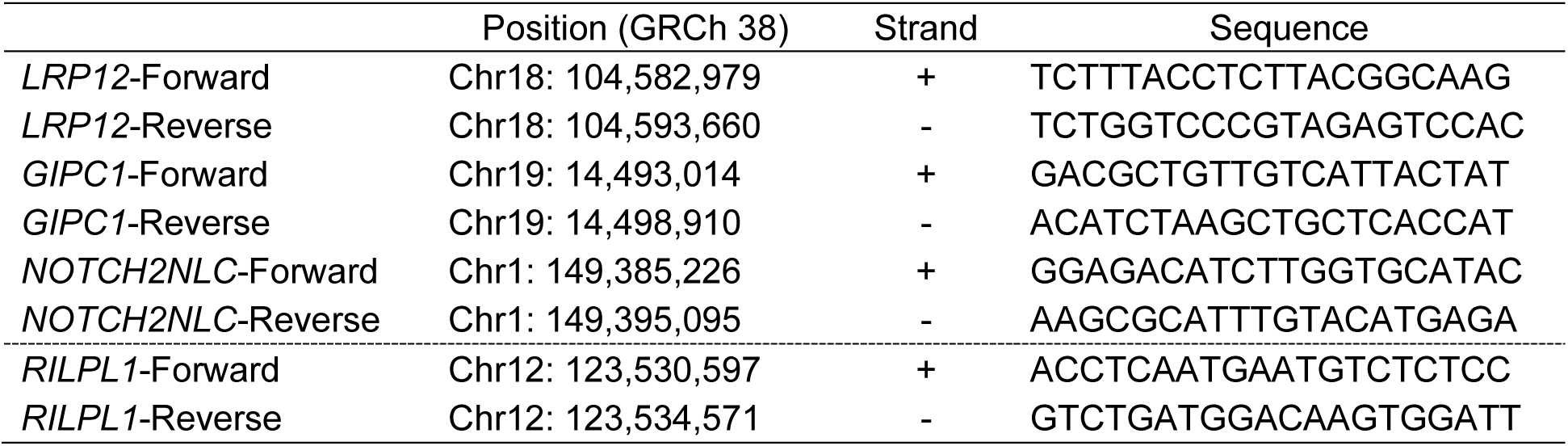
Guide RNA sequences for analysis of four causative genes by nCATS.

